# Striatal dopamine tone is positively associated with body mass index in humans as determined by PET using dual dopamine type-2 receptor antagonist tracers

**DOI:** 10.1101/2023.09.27.23296169

**Authors:** Valerie L. Darcey, Juen Guo, Meible Chi, Stephanie T. Chung, Amber B. Courville, Isabelle Gallagher, Peter Herscovitch, Rebecca Howard, Melissa LaNoire, Lauren Milley, Alex Schick, Michael Stagliano, Sara Turner, Nicholas Urbanski, Shanna Yang, Eunha Yim, Nan Zhai, Megan S. Zhou, Kevin D. Hall

## Abstract

The relationship between adiposity and dopamine type-2 receptor binding potential (D2BP) in the human brain has been repeatedly studied for >20 years with highly discrepant results, likely due to variable methodologies and differing study populations. We conducted a controlled inpatient feeding study to measure D2BP in the striatum using positron emission tomography with both [^18^F]fallypride and [^11^C]raclopride in pseudo-random order in 54 young adults with a wide range of body mass index (BMI 20-44 kg/m^2^). Within-subject D2BP measurements using the two tracers were moderately correlated (r=0.47, p<0.001). D2BP was negatively correlated with BMI as measured by [^11^C]raclopride (r= -0.51; p<0.0001) but not [^18^F]fallypride (r=-0.01; p=0.92) and these correlation coefficients were significantly different from each other (p<0.001). Given that [^18^F]fallypride has greater binding affinity to dopamine type-2 receptors than [^11^C]raclopride, which is more easily displaced by endogenous dopamine, our results suggest that adiposity is positively associated with increased striatal dopamine tone.

## INTRODUCTION

Over 20 years ago, Wang et al. published a seminal paper reporting a negative correlation between body mass index (BMI) in people with obesity and brain dopamine type-2 receptor binding potential (D2BP) measured in the striatum using positron emission tomography (PET) with the radiotracer antagonist [^11^C]raclopride (1). This finding was interpreted as a decrease in dopamine type-2 receptor number with increased adiposity and suggested that people with obesity have a deficiency in dopamine signaling, thereby sharing neurobehavioral characteristics with people suffering from addiction and compulsive behaviors. Similar results of decreased dopamine type-2 receptors were subsequently reported in a seminal rodent study of diet-induced obesity demonstrating addiction-like reward deficits and compulsive eating (2). To date, these studies have been cited very widely and have cemented the idea that obesity is linked to a decrease in striatal dopamine D2 receptors. However, subsequent studies in humans have yielded conflicting results, with some studies showing positive associations between adiposity and D2 receptors or D2BP (3–6), others showing negative associations (\(7, 8), and some showing no association at all ((9–12) (as reviewed by (13). Similarly, several rodent studies have yielded discrepant findings, with some demonstrating that in diet induced obesity, some aspects of D2 receptor biology either decrease (14, 15), increase (16, 17), or remain similar to control animals (15, 18).

Considering the inconsistent findings in humans, some researchers have recently concluded that there is no meaningful relationship between brain dopamine and obesity (19). Alternatively, differences in methodology may explain discrepancies between the studies. For example, in the human studies, different radiotracers with different kinetics have been used and PET scans were conducted at different times of the day in participants that were in different physiological states (e.g., fed or fasted), if at all reported. Only one previous study (20, 21) controlled food intake during the days prior to the PET scan, which was recently shown to affect striatal D2BP in people with obesity (22). Additionally, variations in study populations under investigation – specifically, greater age (6, 23), single sex (24, 25), and limited BMI ranges (3, 12) - may have contributed to the differing results.

A novel hypothesis aimed at explaining the seemingly discrepant human results was proposed by Horstmann et al. who suggested that differences in D2BP between BMI categories are better interpreted as differences in tonic dopamine levels that compete for D2 receptor binding with radiotracers used to measure D2BP (26). They propose that dopamine tone may decrease as people move from low to moderate BMI thereby allowing more tracer to bind with D2 receptors, which is reflected as an increase in D2BP with BMI. In theory, such a model suggests that as people gain weight and move into moderate BMI range, they might experience heightened reward sensitivity due to an amplified phasic dopamine response on a background of low tonic dopamine. In the moderate BMI range, tonic dopamine levels stop decreasing thereby resulting in a flat part of the D2BP vs BMI curve. However, as BMI further increases, dopamine tone is hypothesized to increase and displace the radiotracer from the D2 receptor thereby decreasing D2BP – a state theoretically coupled with a relatively blunted magnitude of phasic dopamine response and decrease in reward sensitivity. Thus, Horstmann et al. hypothesized that the relationship between D2BP and BMI is a curve with a negative quadratic coefficient. Past studies could potentially be reconciled by noting that each study included subjects from a relatively narrow BMI range and therefore sampled from only a small part of the nonlinear curve relating D2BP with BMI and resulting in conflicting linear relationships (26).

To test the neurochemical component of this theory and avoid potentially confounding factors in previous studies, the main objectives of our preregistered study was to measure striatal D2BP using two common radiotracers under highly standardized conditions in adults aged 18-45 years encompassing a wide range of BMIs. Specifically, the participants were admitted as inpatients to the NIH Clinical Center and we measured striatal D2BP using both [^18^F]fallypride and [^11^C]raclopride in pseudorandom order on separate days in the overnight fasted state following a period of controlled dietary stabilization. The primary aims of the study were to detect both quadratic and linear relationships between striatal D2BP and BMI as well as the within subject correlation between D2BP assessed by [^18^F]fallypride and [^11^C]raclopride.

## RESULTS

Sixty-one weight stable adults across a wide BMI range completed an in-patient admission to the NIH Clinical Center to ensure standardized composition and timing of meals as well as compliance with overnight fasting prior to PET scanning (**Table 1; Supplemental Figure 1**). Participants were asked to completely consume the provided eucaloric standard diet (50% calories from carbohydrate, 35% from fat, 15% from protein) for 3-5 days in advance of an inpatient admission where they continued the standardized diet for 5 additional days (**Figure 1**). During their inpatient admission, participants completed PET neuroimaging in the overnight fasted state with both [^11^C]raclopride and [^18^F]fallypride in pseudorandom order in addition to a high-resolution neuroanatomical magnetic resonance image (MRI).

**Figure 1.**
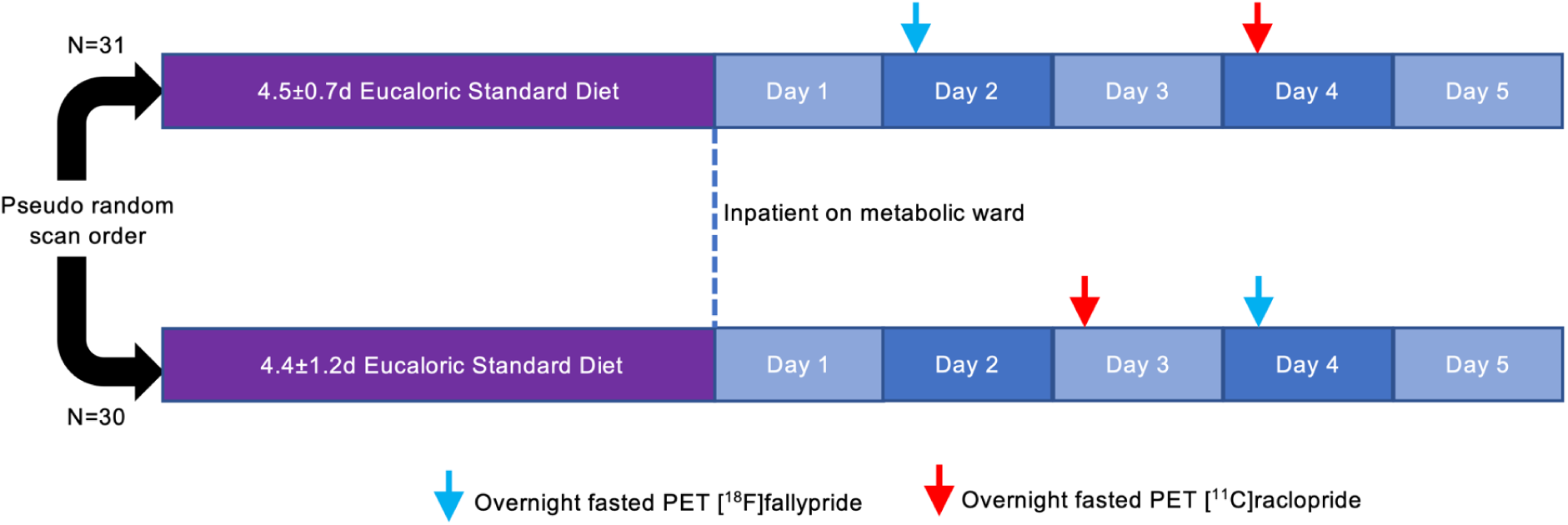
Study design. Sixty-one men and women consumed a provided weight-stabilizing standardized diet for an average of 4.5±1.0 days prior to admission to the NIH Clinical Center for testing. During their inpatient stay, participants continued their dietary stabilization. Between their second day of admission and the morning of their discharge, participants completed morning PET scans in the overnight fasted state with [^18^F]fallypride and [^11^C]raclopride on separate days in pseudo-random order after an average of 6.5±1.3 and 6.8±1.1 total days of dietary stabilization, respectively. Arrows indicate mode value for scan completion day.

**Table 1.** Characteristics of participants completing PET scanning with technically adequate. Means and standard deviations indicated.

|  | Enrolled participants | [18F] fallypride | [11C] raclopride | [11C] raclopride & [18F] fallypride |
| --- | --- | --- | --- | --- |
| N | 61 | 57 | 56 | 54 |
| Females | 40 (65%) | 38 (66.7%) | 36 (64.3%) | 35 (64.8%) |
| Age (years) | 31.7 ± 7.3 | 31.8 ± 7.2 | 31.6 ± 7.1 | 31.4 ± 7.1 |
| Race |  |  |  |  |
| Black | 32 (52.5%) | 28 (49.1%) | 30 (53.6%) | 28 (51.9%) |
| White | 18 (29.5%) | 18 (31.6%) | 17 (30.4%) | 17 (31.5%) |
| Asian | 7 (11.5%) | 7 (12.3%) | 6 (10.7%) | 6 (11.1%) |
| Other/Multiple | 4 (6.6%) | 4 (7.1%) | 3 (5.4%) | 3 (5.6%) |
| Body weight (kg) |  |  |  |  |
| Mean | 85.9 ± 25.3 | 84.6 ± 24.2 | 85.0 ± 24.2 | 84.4 ± 23.6 |
| Range | 45.9 – 148.6 | 45.9 – 148.6 | 45.9 – 148.6 | 45.9 – 148.6 |
| Body fat (%) |  |  |  |  |
| Mean | 35.0 ± 12.6 | 34.7 ± 12.1 | 34.4 ± 12.0 | 34.1 ± 12.1 |
| Range | 11.3 – 59.0 | 11.3 – 56.7 | 11.3 – 52.4 | 11.3 – 52.4 |
| BMI (kg/m<sup>2</sup>) |  |  |  |  |
| Mean | 30.1 ± 8.2 | 29.6 ± 7.6 | 29.7 ± 7.7 | 29.5 ± 7.5 |
| Range | 20.3 – 52.8 | 20.3 – 44.4 | 20.3 – 44.8 | 20.3 – 44.4 |

### Within subject correlation between D2BP measured by [^11^C]raclopride and [^18^F]fallypride

Both D2BP_raclo_ and D2BP_fally_ were measured during a period of dietary stabilization and after confirmed overnight fasts of similar duration. Across all participants with both scans meeting quality control requirements (n=54), D2BP_raclo_ and D2BP_fally_ were positively correlated within the region of interest (ROI) defining the whole striatum (r=0.468, p<0.001) (**Figure 2A**). ROI analyses within bilateral striatal structures including caudate, putamen, accumbens and pallidum revealed significant within-subject positive correlations between D2BP_raclo_ and D2BP_fally_ in caudate (r=0.467, p<0.001), putamen (r=0.547, p<0.001), accumbens (r=0.484, p<0.001) and pallidum (r=0.685, p<0.001) (**Supplementary** Figure 2).

**Figure 2.**
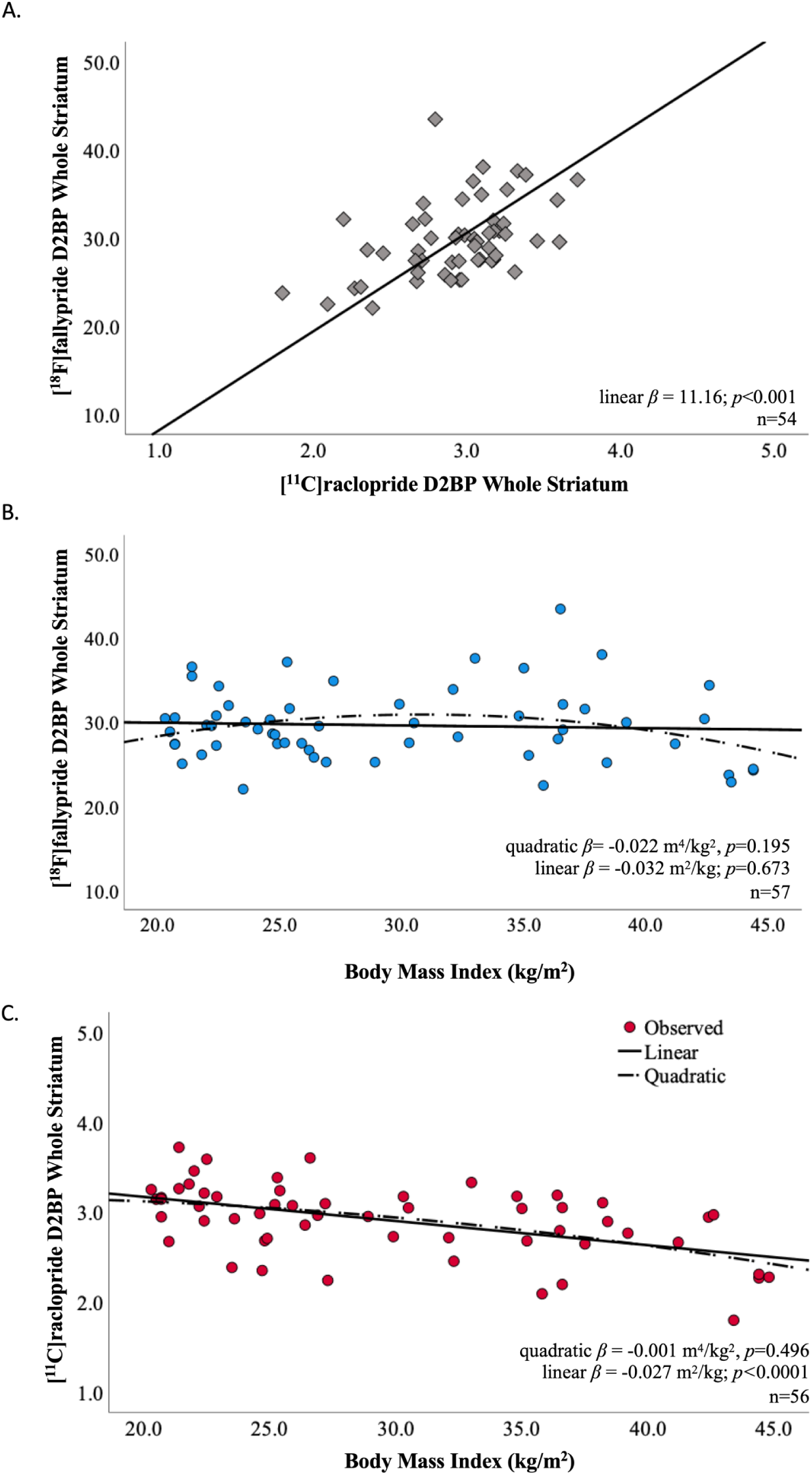
Striatal D2BP as measured within subject by [^18^F]fallypride and [^11^C]raclopride were correlated but differentially related to BMI. (A) Within-subject measurements of D2BP using [^18^F]fallypride and [^11^C]raclopride in the overnight fasted state were correlated using the whole striatum in a region of interest analysis. Trendline and slope parameters reflect standard major axis regression. (B) Whole striatal region of interest analysis using [^18^F]fallypride indicated no significant relationship between D2BP and BMI. (C) Whole striatal region of interest analysis using [^11^C]raclopride indicated a negative linear relationship between BMI and D2BP, but no significant quadratic relationship.

Voxelwise analyses within the striatal mask supported the ROI results, revealing clusters where D2BP_raclo_ and D2BP_fally_ were positively correlated in regions spanning dorsal and ventral striatum (**Supplementary** Figure 3; **Supplementary Table 1**).

### BMI was not related to striatal D2BP measured with [^18^F]fallypride but was negatively linearly related to striatal D2BP measured with [^11^C]raclopride

The mean whole striatal D2BP as measured by [^18^F]fallypride was neither quadratically (p=0.195) nor linearly (p=0.673) associated with BMI (**Figure 2B**). These results were robust to the exclusion of data from one participant whose striatal D2BP measured by [^18^F]fallypride was determined to be an outlier (27) (**Supplementary** Figure 4). In contrast, mean whole striatal D2BP measured using [^11^C]raclopride decreased linearly with increasing BMI (r= -0.514, p<0.0001) but was not quadratically associated with BMI (p=0.496) in the same participants under the same scanning conditions (**Figure 2C**). No outliers were detected for striatal D2BP measured using [^11^C]raclopride.

Within striatal sub-regions of interest, BMI was not significantly related to D2BP as measured by [^18^F]fallypride (**Figure 3, left column**). Age was negatively correlated with D2BP_fally_ across the whole striatum (r=-0.341, p=0.009), but adjustment for age and sex did not impact the lack of relationship between [^18^F]fallypride D2BP and adiposity using either BMI (**Supplementary** Figure 5**, left column**) or percent body fat (**Supplementary** Figures 6 **& 7, left columns**). Significant negative linear relationships between BMI and D2BP_raclo_ were observed in all striatal subregions of interest except the pallidum (**Figure 3, right column**), and the significance of the relationships across the striatum as a whole and dorsal striatum (caudate and putamen) persisted after adjustment for age and sex (**Supplementary** Figure 5**, right column**). D2BP_raclo_ across the whole striatum was negatively related to age (r=-0.451, p<0.001). Percent body fat was negatively related to D2BP_raclo_ within the whole striatum (r=-0.337, p=0.011) and putamen (r=-0.367, p=0.005), uncorrected for multiple comparisons (**Supplementary** Figure 6**, right column**) but significance did not survive adjustment for age and sex (**Supplementary** Figure 7**, right column**).

**Figure 3.**
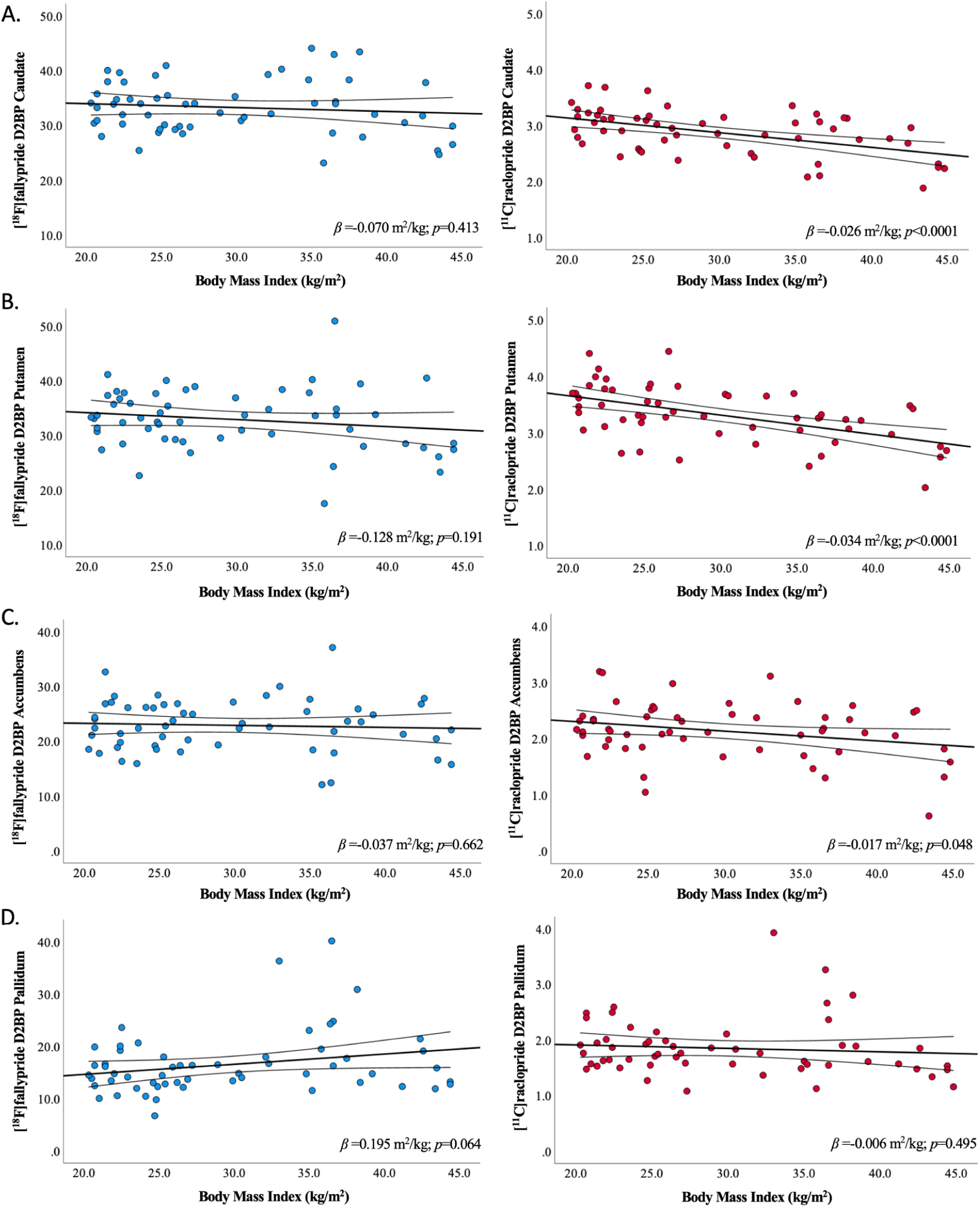
Relationship between BMI and D2BP as measured by [^18^F]fallypride and [^11^C]raclopride across striatal regions of interest. Bilateral striatal region of interest analyses reflecting relationships between BMI and D2BP as measured by [^18^F]fallypride (blue circles, left column; n=57) and [^11^C]raclopride (red circles, right column; n=56) in (A) caudate, (B) putamen, (C) accumbens, and (D) pallidum. Regression line and 95%CI indicated. ROI analyses are uncorrected for multiple comparisons.

Voxelwise analyses of D2BP were restricted to within the striatal region mask (small volume correction) and revealed that D2BP as measured by [^18^F]fallypride was positively related to BMI in two clusters (proximal to right and left caudate) and negatively related to BMI in the right thalamus. However, only the cluster proximal to the right caudate marginally survived correction for multiple comparisons using AFNI’s 3DClustSim (**Supplementary** Figure 8**; Supplementary Table 1**).

Voxelwise analyses of D2BP within the striatal mask using [^11^C]raclopride supported the ROI analyses, revealing clusters with peaks where D2BP negatively correlates with BMI in the left and right putamen surviving correction for multiple comparisons (**Supplementary** Figure 9**; Supplementary Table 1**).

### The linear relationship between D2BP_raclo_ and BMI is significantly more negative than between D2BP_fally_ and BMI

A statistical comparison of standardized correlation coefficients of D2BP_raclo_ with BMI and D2BP_fally_ with BMI showed that the linear relationship between D2BP_raclo_ and BMI was significantly more negative than that of the D2BP_fally_ relationship with BMI (**Table 2**). Voxelwise maps reflecting the difference in correlation coefficients between the D2BP_raclo_× BMI correlation and D2BP_fally_× BMI correlation produced a cluster in the left caudate-putamen boundary which survived multiple comparisons correction, suggesting this region has the greatest difference in correlation coefficients (**Supplementary** Figure 10**; Supplementary Table 1**).

**Table 2.** Standardized correlation coefficients between D2BP and measure of adiposity across regions of interest for n=54 participants with both radiotracer scans available. D2BP and adiposity measures normalized to facilitate comparison between tracers (Corresponding figures located in Supplemental Figures (5-7).

|  | *D2BP<sub>fally</sub>* | *p* | *D2BP<sub>raclo</sub>* | *p* | *p (D2BP<sub>fally</sub> vs. D2BP<sub>raclo</sub>)* |
| --- | --- | --- | --- | --- | --- |
| *BMI* |  |  |  |  |  |
| Whole striatum | -0.013 | 0.923 | -0.509 | <0.001 | <0.001 |
| Caudate | -0.066 | 0.635 | -0.482 | <0.001 | 0.002 |
| Putamen | -0.132 | 0.343 | -0.536 | <0.001 | 0.001 |
| Accumbens | 0.000 | 0.999 | -0.238 | 0.083 | 0.091 |
| Pallidum | 0.245 | 0.074 | -0.056 | 0.687 | 0.007 |
| *Percent Body Fat* |  |  |  |  |  |
| Whole striatum | -0.056 | 0.688 | -0.314 | 0.021 | 0.068 |
| Caudate | -0.026 | 0.851 | -0.192 | 0.164 | 0.247 |
| Putamen | -0.131 | 0.345 | -0.350 | 0.010 | 0.089 |
| Accumbens | -0.072 | 0.602 | -0.182 | 0.188 | 0.437 |
| Pallidum | 0.039 | 0.778 | -0.131 | 0.345 | 0.125 |
| **Adjusted for age and sex** |  |  |  |  |  |
|  | *D2BP<sub>fally</sub>* | *p* | *D2BP<sub>raclo</sub>* | *p* | *p (D2BP<sub>fally</sub> vs. D2BP<sub>raclo</sub>)* |
| *BMI* |  |  |  |  |  |
| Whole striatum | 0.141 | 0.309 | -0.337 | 0.013 | 0.001 |
| Caudate | 0.093 | 0.503 | -0.326 | 0.016 | 0.004 |
| Putamen | 0.002 | 0.987 | -0.382 | 0.004 | 0.003 |
| Accumbens | 0.095 | 0.497 | -0.162 | 0.241 | 0.071 |
| Pallidum | 0.259 | 0.059 | 0.055 | 0.692 | 0.063 |
| *Percent Body Fat* |  |  |  |  |  |
| Whole striatum | 0.104 | 0.453 | -0.188 | 0.174 | 0.043 |
| Caudate | 0.119 | 0.393 | -0.155 | 0.262 | 0.058 |
| Putamen | 0.022 | 0.873 | -0.222 | 0.106 | 0.065 |
| Accumbens | 0.075 | 0.590 | -0.047 | 0.735 | 0.391 |
| Pallidum | 0.139 | 0.316 | 0.025 | 0.859 | 0.302 |

## DISCUSSION

Under tightly controlled experimental conditions, we observed a negative linear relationship between BMI and striatal D2BP measured using [^11^C]raclopride in adults across a broad range of adiposity. These findings align with the influential study of Wang and colleagues who also used [^11^C]raclopride to measure D2BP (1). However, there were no significant associations between BMI and striatal D2BP determined by [^18^F]fallypride despite moderate within-subject correlations between the [^11^C]raclopride and [^18^F]fallypride measurements of striatal D2BP. Furthermore, the correlation coefficients between BMI and striatal D2BP measured using the two tracers were significantly different. These results held true even when considering alternative measures of adiposity, such as percent body fat, and when adjusting D2BP for factors like sex and age.

Given that both [^11^C]raclopride and [^18^F]fallypride PET scans were conducted on the same inpatient subjects in matched physiological states, our findings suggest that the use of different radiotracers may partly explain the inconsistent results over the last two decades concerning the relationship between striatal D2BP and adiposity in humans (28). The differing kinetics of [^11^C]raclopride and [^18^F]fallypride could potentially explain the discordant associations between adiposity and striatal D2BP and may shed light on striatal dopamine physiology and its connection to human obesity.

For instance, our results might be explained by considering the binding kinetics of [^11^C]raclopride and [^18^F]fallypride for dopamine D2 and D3 receptors. Notably, [^11^C]raclopride binds relatively similarly to both D2 and D3 receptors whereas [^18^F]fallypride has a higher selectivity for D2 receptors (29). Thus, the observed lack of correlation between BMI and striatal D2BP measured with [^18^F]fallypride, as opposed to the negative correlation seen with [^11^C]raclopride, could potentially stem from the presence of fewer D3 receptors along with a relatively consistent number of D2 receptors with increasing BMI. However, this explanation faces two challenges. First, D2 receptors are more abundant than D3 receptors within the striatum (30),making it unlikely for a decrease in striatal D3 receptors with BMI to be detected by [^11^C]raclopride. Second, such an effect would likely be most apparent in the ventral striatum, where D3 receptors are relatively more abundant compared to the dorsal striatum (31). However, the opposite was true, with the negative correlation between BMI and D2BP as measured by [^11^C]raclopride being present in the dorsal striatum, a region with minimal D3 receptors in comparison to D2 receptors (32).

We believe that the most likely interpretation of our data is that adiposity is associated with increased dopamine tone and this relationship may be particularly prominent in the dorsal striatum. Compared to [^18^F]fallypride, the [^11^C]raclopride tracer has a lower affinity for the D2 receptor making it more easily displaced by endogenous dopamine (33). Thus, increased dopamine tone at higher levels of adiposity would be expected to result in a more negative correlation between adiposity and D2BP as measured by [^11^C]raclopride as compared with [^18^F]fallypride. While this is indeed what we observed, it is important to note that our study lacks direct measures of dopamine tone.

The absence of a significant association between adiposity and striatal D2BP measured with [^18^F]fallypride does not necessarily imply that D2 receptor number is unrelated to adiposity. Despite its high affinity for the D2 receptor, [^18^F]fallypride is somewhat displaceable by manipulations which may increase endogenous dopamine (22, 34, 35). Therefore, we cannot definitively rule out a positive association between adiposity and striatal D2 receptor number because increased dopamine tone at higher levels of adiposity might mask this relationship by displacing [^18^F]fallypride and result in a more shallow slope not significantly different from zero. Nevertheless, a previous study using a D2 receptor-specific radiotracer not displaceable by endogenous dopamine found no relationship between D2BP and adiposity (9).

We did not find supporting evidence for Horstman et al.’s hypothesis that the relationship between striatal D2BP and BMI is quadratic (28). However, our data support their suggestion that obesity is associated with increased striatal dopamine tone, but the mechanisms remain uncertain. Tonic dopamine levels are proportional to the number of active DA terminals and spontaneous, irregular single-spike firing activity of midbrain DA neurons (36, 37). There is early indication that capacity to synthesize dopamine may be blunted in obesity (38–40). However, it may be that a limited synthetic capacity impairs the ability to mount a phasic dopamine response to reward-predicting stimuli as opposed to limiting basal dopamine tone in humans (41, 42), particularly in the context of obesity. It is intriguing to speculate that an increase in spontaneous activity of dopamine neurons or number of active terminals may be due proximally to alterations in inhibitory influence from GABAergic inputs (43), or various hormones like ghrelin, leptin, and insulin (44–46).

Our study was not designed to elucidate mechanisms, and its cross-sectional design prevents us from determining whether increased adiposity leads to heightened dopamine tone or the reverse. Nevertheless, increased striatal dopamine tone in people with obesity may increase incentive salience and wanting of rewarding stimuli (47) while simultaneously blunting the phasic dopamine response that has been associated with salient events and the subjective experience of rewards (48).This potential effect may be compounded by tonic dopaminergic modulation of prefrontal cortical synaptic afferents that can attenuate behavioral control and goal-directed information processing (49). Hence, our findings could align with the “reward deficiency hypothesis”, which proposes that people with obesity experience greater wanting for palatable foods, yet their consumption is not accompanied by the expected reward thereby promoting overconsumption (50).

However, the role of dopamine in the control of food intake and regulation of body weight is likely to be much more complicated than the reward deficiency hypothesis suggests. Studies in rodents have identified specific neuron populations in the hypothalamus that control homeostatic food intake that has long been known to influence the brain’s reward system, possibly by augmenting dopamine signaling in the brain’s reward circuitry (51, 52). Indeed, food restriction increases motivation, incentive salience, and susceptibility to addiction in experimental models (53). Furthermore, recent studies in mice suggest that brain reward pathways involving dopamine interact with hypothalamic neurons responsible for homeostatic feeding, forming a bidirectional circuit (54). Exposure of mice to energy-dense and palatable diets affects this bidirectional circuit and leads to excessive food consumption and the development of obesity, while devaluing a non-obesogenic chow diet (55). Thus, it is possible that alterations in striatal dopamine tone may affect both hedonic and homeostatic control of feeding and may thereby contribute to excess adiposity in people with obesity.

## AUTHOR CONTRIBUTIONS

VLD, PH and KDH designed the research study. ABC, ST, SY, and STC contributed to research design, data collection and analysis. MC, IG, RH, ML, LM, AS, MSS, EY, NZ, MSZ conducted experiments and collected data. VLD and JG analyzed data and performed statistical analysis. VLD and KDH drafted the manuscript. All authors contributed intellectually and approved the manuscript.

## Data Availability

Data reported in the present study are available upon reasonable request to the authors

## ACKNOWLEDGEMENTS

This work was supported by the Intramural Research Program of the National Institutes of Health, National Institute of Diabetes and Digestive and Kidney Diseases and by the NIH Center on Compulsive Behaviors via the NIH Shared Resource Subcommittee. We thank the PET Department staff and technologists, nursing and nutrition staff at the NIH Metabolic Clinical Research Unit for their invaluable assistance with this study. We thank David Rosenberg for his computer simulations of D2BP correlations as determined by [^11^C]raclopride and [^18^F]fallypride that were used in our power calculations and Nathalie Ginovart, Anthony Grace, and Gene Jack Wang for their helpful comments on our results. We are most thankful to the study subjects who volunteered to participate in this demanding protocol.

## METHODS

Sixty-one adults provided informed consent to participate in a dual PET radiotracer study investigating the relationship between D2R availability and BMI under controlled dietary conditions (ClinicalTrials.gov NCT03648892). Participants were recruited from the community over a wide BMI range and approximately evenly sampled in each of three BMI categories (18.5 kg/m^2^ ≤ BMI < 25 kg/m^2^, 25 kg/m^2^ ≤ BMI < 35 kg/m^2^, BMI ≥ 35 kg/m^2^) to ensure sufficient BMI range to test the quadratic hypothesis. Eligible volunteers were English-speaking, weight stable (less than ± 5% change in the past month), between 18-45 years of age, BMI ≥18.5 kg/m^2^. They had no history of bariatric surgery, metabolic disorders, previous traumatic head injury or neurological disorders, severe food allergies (e.g., dairy, gluten) impaired activities of daily living, high blood pressure (>140/90 mm Hg), or current use of medication influencing metabolism or psychiatric medications. They did not have psychiatric conditions or disordered eating (EDE-Q, DSM Cross Cutting Symptom Measure Self Rated Level 1), nicotine dependence, drug use or in past 12 months (confirmed via urine toxicology at screening visit), binge drinking over previous 6 months, excessive caffeine consumption, or safety contraindications to MRI. Females were excluded if they were pregnant or lactating. In the women reporting regular menses (not using hormonal contraceptives) (n=31), inpatient admissions started on day 17.4±9.9 of their cycle. Participants self-identified race and ethnicity at the time of admission to the NIH Clinical Center. Handedness was not exclusionary. Participants completed the 10-item Edinburgh Handedness questionnaire to determine laterality quotient (56) and 96.7% of participants (n=59) were determined to be right-handed (laterality quotient >0).

### Method Details

This study was conducted between September 26, 2018 and February 17, 2023. During their inpatient admission, participants completed two PET sessions in pseudorandom order after a confirmed overnight fast. On average, [^18^F]fallypride scans and [^11^C]raclopride scans were completed after 6.5±1.3 and 6.8±1.1 total days of dietary stabilization, respectively. MRI was completed to collect high resolution T-1 weighted structural brain images on which to register individual subject PET data.

The CONSORT diagram provides details of enrollment (**Supplementary** Figure 1). No participants withdrew from the inpatient portion after enrollment. Thirty participants received the order of [^11^C]raclopride followed by [^18^F]fallypride while 31 received the order of [^18^F]fallypride followed by [^11^C]raclopride. The initial radiotracer order randomization schedule was provided to the PET Department and was accommodated within the radiotracer production schedule when possible. Of 61 enrolled participants, [^11^C]raclopride scan data are available for n=56 (n=1 participant declined, n=2 scans not performed due to tracer production issue, n=2 scans completed but did not pass quality control on time activity curves) and [^18^F]fallypride scan data are available for n=57 (n=4 scans completed but did not pass quality control on time activity curves). Full PET data [^11^C]raclopride and [^18^F]fallypride) are available on n=54 participants (**Table 1**). All participants completed structural MRI. All study procedures were approved by the Institutional Review Board of the National Institute of Diabetes & Digestive & Kidney Diseases and the NIH Radiation Safety Committee; participants were compensated for their participation.

### Anthropometrics

Height was measured in centimeters using a wall stadiometer (Seca 242, Hanover, MD, USA) and weight was measured in kilograms using a digital scale (Scale-Tronix 5702, Carol Steam, IL, USA). All measurements were obtained after an overnight fast while participants were wearing comfortable clothing.

### Body Composition

During the inpatient stay, participants each completed one Dual Energy X-Ray Absorptiometry (DEXA) scan while wearing hospital gown/scrubs to determine body composition (General Electric Lunar iDXA; General Electric; Milwaukee, WI, USA).

### Metabolic Diet

Participants were placed on a standard eucaloric diet (50% carbohydrate, 15% protein, 35% fat) with daily energy needs calculated using the Mifflin-St Jeor equation and standard activity factor of 1.5. All meals were prepared in the NIH Clinical Center Nutrition Department Metabolic Kitchen with all foods and beverages weighed on a gram scale (Mettler Toledo Model MS12001L/03).

For the run-in phase, participants were provided with 3-5 days of meals for retrieval from the NIH Clinical Center and consumed them at home prior to admission. Participants were instructed to consume all foods and beverages provided. Any food or beverage not consumed was returned and weighed back. Participants were also instructed to continue their usual caffeine intake in calorie-free forms (e.g., black coffee, diet soda) and abstain from alcohol during this period. For any foods or beverages participants consumed that were not part of the standardized run-in diet, participants were asked to provide a description and amount of what was consumed so that total daily nutrient intake was captured. The eucaloric standardized outpatient diet was provided for an average of 4.5±1.0 days (range 0 – 5 days). Due to COVID-19 pandemic precautions, one participant was admitted without having completed a diet stabilization, and 3 participants completed some or all of their 3-5 day diet stabilization in the inpatient setting. The remainder (n=57) consumed their stabilization diet as outpatients.

During the inpatient phase, participants continued the same diet and were instructed to consume all foods and beverages provided. All subjects were confined to the metabolic ward throughout their inpatient stay without access to outside food. Meals were consumed under observation. Any uneaten food was weighed back and energy and macronutrients were replaced at the next available meal as needed. Diets were designed using ProNutra software (version 3., Viocare, Inc.). No adverse events, harms or unintended effects resulted from provision of standardized eucaloric diet.

### Magnetic Resonance Imaging

During their inpatient stay, high resolution anatomical brain MRI was acquired for each subject. Due to the duration of data collection, extended by the COVID-19 pandemic, T1 weighted structural MRIs were collected on 3T Siemens Verio (n=21; TE = 2.98 ms, TR = 2.3 ms, TI = 900 ms, flip angle 9°, slice thickness = 1.2 mm, voxel size 1*1*1.2mm), and on 3T GE MR-750 Discovery scanner (n=6, TE = 3.04 ms, TR = 7.648 ms, TI = 1060 ms, flip angle 8°, slice thickness = 1.0 mm, voxel size 1*1*1mm; n=32, TE= 3.46 ms, TR = 8.156 ms, TI = 900 ms, flip angle 7°, slice thickness = 1.0 mm, voxel size 1*1*1 mm) for each subject. Quality of individual subject data were checked by study team [VLD & JG].

The anatomical images were parcellated with FreeSurfer software to generate ROI binary mask volumes in each subject in the putamen, caudate, accumbens, pallidum, and the cerebellum (reference region) (http://surfer.nmr.mgh.harvard.edu). All individual ROI masks were visually checked.

### Positron Emission Tomography

All PET scanning was performed using a High Resolution Research Tomograph (HRRT), (Siemens Healthcare, Malvern, PA), a dedicated brain PET scanner with resolution of 2.5 - 3.0 mm and a 25 cm axial field of view. Transmission scanning was performed with a ^137^Cs rotating point source scan to correct for attenuation. After an overnight fast matched in duration within subject, a bolus of either approximately 5 mCi of [^18^F]fallypride or 20 mCi of [^11^C]raclopride was infused intravenously using a Harvard® pump in semirandom order as discussed above.

The molar activity of [^18^F]fallypride was approximately 7459 mCi/µmol and the radiochemical purity of the radiotracer was > 90%. PET emission data for [^18^F]fallypride were collected starting at radiotracer injection over 3.5 h, in three blocks separated by two 10-min breaks. Thirty-three frames were acquired in list mode at times 0, 0.25, 0.5, 0.75, 1, 1.25, 1.5, 1.75, 2, 2.5, 3, 3.5, 4, 4.5, 5, 6, 7, 8, 9, 10, 12.5, 15, 20, 25, 30, 40, 50, 60, 90, 110, 130, 170, 200 min. The molar activity of [^11^C]raclopride was approximately 4865 mCi/µmol and the radiochemical purity of the radiotracer was >90%. PET emission data for [^11^C]raclopride were collected starting at radiotracer injection over one block lasting 75 minutes. Twenty-four frames were acquired in list mode at times 0, 0.5, 1, 1.5, 2.0, 2.5, 3, 4, 5, 6, 8, 10, 15, 20, 25, 30, 35, 40, 45, 50, 55, 60, 65, 70 min. During each scan block, the room was illuminated and quiet, and each subject was instructed to keep their head as still as possible, relax, and try to avoid falling asleep. The image reconstruction process corrected for head motion which was tracked throughout each scan using an optical head tracking sensor (Polaris Vicra, Northern Digital Inc., Shelburne, VT, USA).

Each scan consisted of 207 slices (slice separation = 1.2 mm). The fields of view were 31.2 cm and 25.2 cm for transverse and axial slices, respectively. The PET images were aligned within each scan block with 6-parameter rigid registration using 7th order polynomial interpolation and each block was aligned to the volume taken at 20 min of the first block. The final alignments were visually checked, with translations varying by <5 mm and the rotations by <5 degrees.

<u>For region of interest analyses,</u> individual participants’ anatomical MRI images were co-registered to the aligned PET images by minimizing a mutual information cost function for each individual participant. Time-activity curves for each tracer concentration in the Freesurfer-generated ROIs were extracted and kinetic parameters were fit to a two-compartment model (with the cerebellum used as the reference tissue given negligible D2/3R specific binding (57) to determine regional D2BP (<u>Lammertsma and Hume 1996</u>).

For voxelwise analyses, each individual’s anatomical MRI was nonlinearly transformed into the Talairach space using AFNI 3dQwarp, and the transformation matrix was applied to the PET images which were then smoothed with a 5-mm full-width, half-max Gaussian kernel. Final coregistration was visually checked. Data were exported from Talairach space to MATLAB where time-activity curves for tracer concentration in each voxel were fit to a kinetic model using the cerebellum as a reference tissue to determine D2BP at each voxel and exported back to Talairaich space for group level spatial analyses.

### Statistics

Power calculations were performed with computer simulation to detect a quadratic relationship between D2BP and BMI with 80% of power and 5% of type I error. Based on the review by Horstmann et al. (28), we assumed a quadratic effect of -0.029 m^4^/kg^2^ and a linear effect of 1.913 m^2^/kg. Equal numbers of BMI’s were randomly drawn from three normal distributions (mean ± SD: 21.75 ± 3.15; 30 ± 4.3; 40 ± 4.3 kg/m^2^) to represent the three BMI strata used in the experimental design. The D2BP value for each simulated subject was calculated from its BMI value plus normally-distributed noise (SD = 4). The parameters for the BMI and noise distributions were derived from our previous study (58). This simulated sample was analyzed using regression analysis and the p-value for the quadratic term was calculated. These simulations were repeated 10000 times and the percentage of p-values less than 0.05 determined the power to detect a significant quadratic effect. Our computer simulation suggested that a minimum of 39 subjects (13 per BMI strata) were required to detect a quadratic relationship between BMI and D2BP. This sample size would also be sufficient to provide 89% power to detect a moderate linear association (r = ±0.45) with a slope of magnitude ≥ 0.25 m^2^/kg between BMI and D2BP in caudate and putamen using [^18^F]fallypride. Finally, this sample size would also provide >80% power to detect a correlation of r > 0.4 between the binding potential of two DA D2 receptor antagonists [^11^C]raclopride and [^18^F]fallypride. Our recruitment exceeded the minimum sample size requirement.

In the ROI analyses, associations between either BMI or percent body fat and D2BP were evaluated with regression analyses. Person correlation coefficients were also reported. Supplementary regression analyses include adiposity (BMI or percent body fat) variables adjusted for sex and age. For associations between D2BP_raclo_ and D2BP_fally_ within ROIs, major axis regressions were also conducted to account for potential measurement error in D2BP calculated from both tracers. Statistical analyses were performed using IBM SPSS Statistics (Version 28.0.1.1, Chicago, IL, USA).

In the voxel-wise analyses, regional clusters where D2BP’s are highly correlated with BMI were identified with regression analysis in AFNI’s 3dttest++ (https://afni.nimh.nih.gov/). Since high D2BP occurs mainly in striatum, small volume corrections were implemented within each hemisphere where D2BP >1.5. A bi-sided uncorrected voxel-wise threshold of p<0.1 was used with a cluster extent minimum of 20 voxels (faces touching). Resultant clusters were deemed to survive correction for multiple comparisons using 3dClustSim at alpha of <0.05 and a threshold of 34 voxels.

### Study Approval

All study procedures were approved by the Institutional Review Board of the National Institute of Diabetes & Digestive & Kidney Diseases and the NIH Radiation Safety Committee. Written informed consent was received prior to participation and compensation was provided.

