## Supplemental Material for "Striatal dopamine tone is positively associated with body mass index in humans as determined by PET using dual dopamine type-2 receptor antagonist tracers"

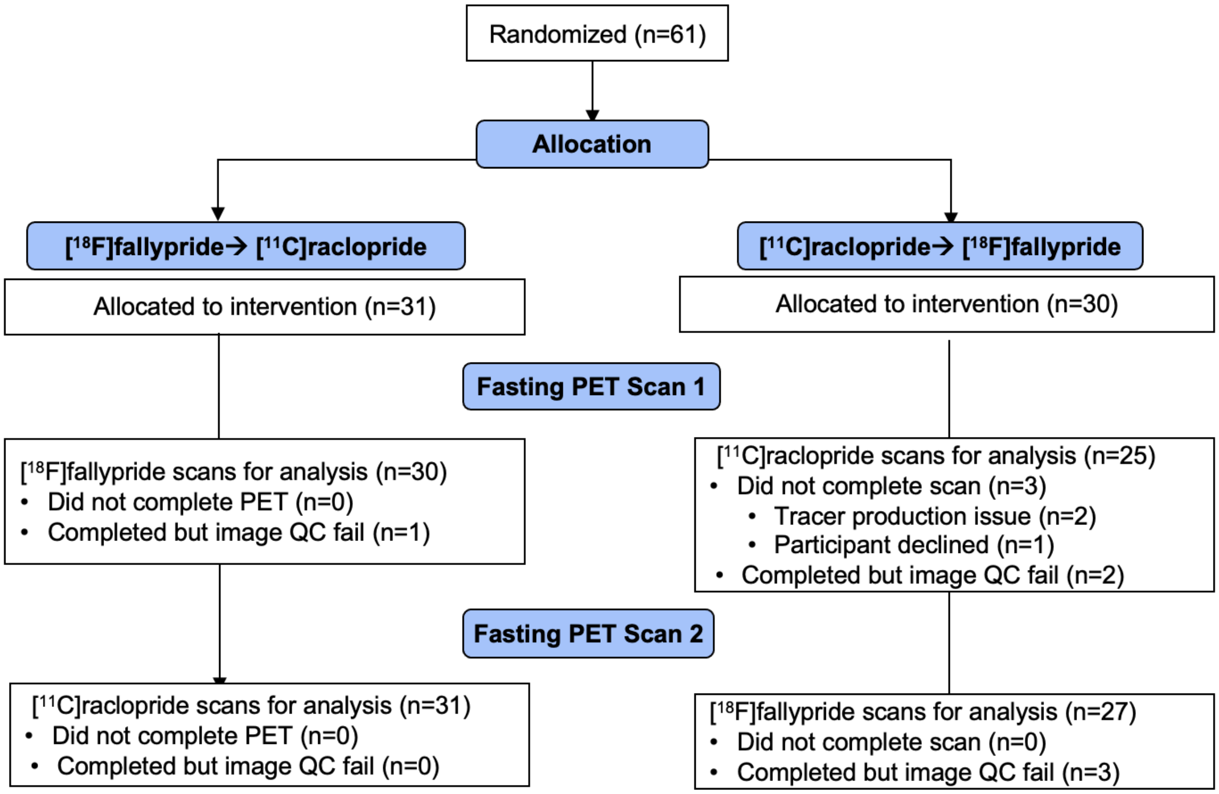


**Supplementary Figure 1.** CONSORT diagram.

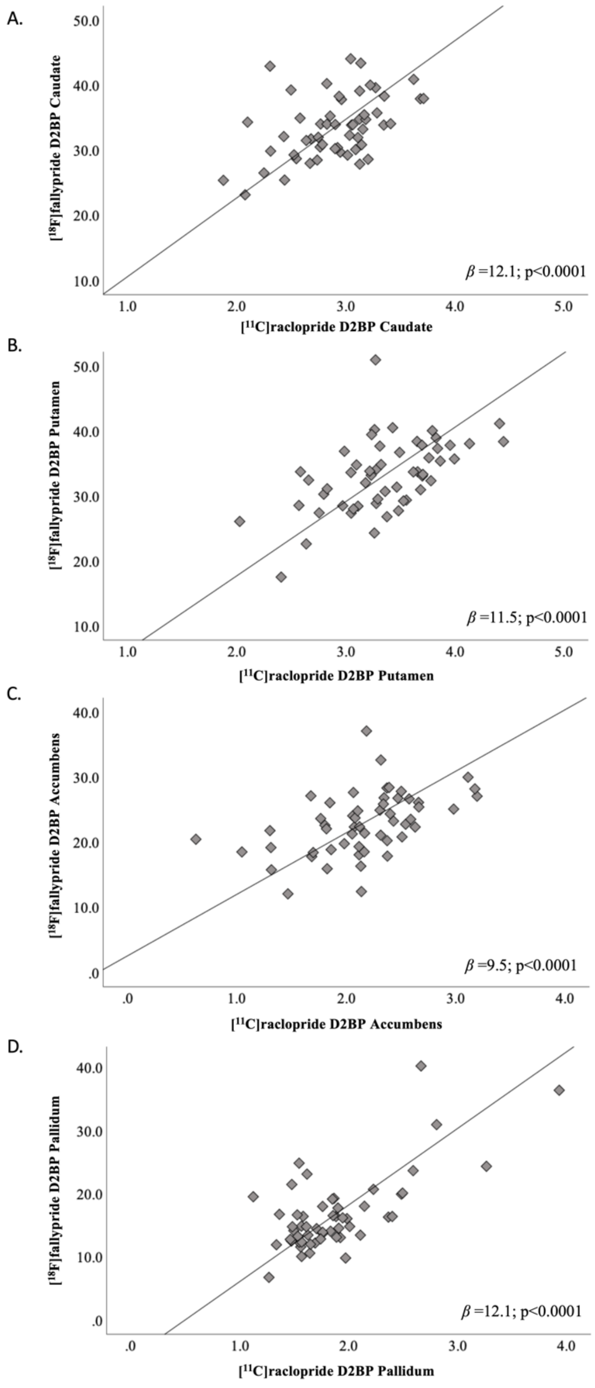


**Supplementary Figure 2. Relationship between $D2BP_{raclo}$ and $D2BP_{fally}$ within bilateral striatal regions of interest.** Striatal regions of interest analyses in n=54 participants with valid scans using both tracers in (A) caudate, (B) putamen, (C) accumbens, and (D) pallidum. Trendline and slope parameters reflect standard major axis regression. Not corrected for multiple comparisons.

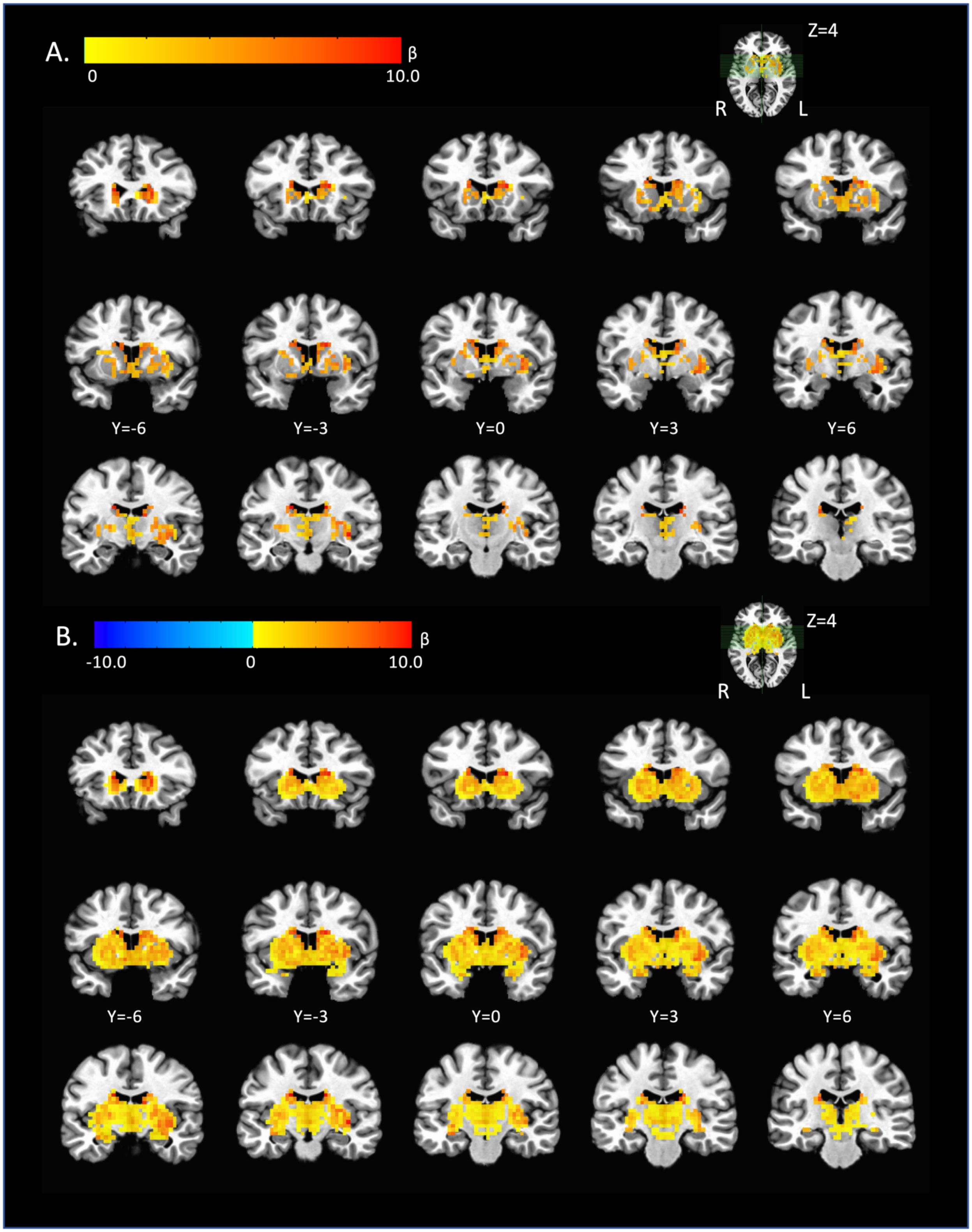


**Supplementary Figure 3. Voxelwise analysis of correlation between $D2BP_{raclo}$ and $D2BP_{fally}$ across n=54 participants. (**A) Thresholded. (B) Unthresholded maps.

**Supplementary Table 1. Locations of striatal clusters with significant correlations.** PET resolution 3.5mm$^3$. Imaging analyses conducted in Analysis of Functional Neuroimaging (AFNI) within striatal region binding potential mask (clusters defined by voxels with faces touching).

| | **Location of peak** | | | **Voxels** | **Size (mm$^3$)** | **t-stat** *(e)* | **alpha** |
|---|---|---|---|---|---|---|---|
| | x | y | z | | | | |
| $D2BP_{fally}$ x $D2BP_{raclo}$ *(a)* | | | | | | | |
| Left caudate, posterior | 15.8 | 11.5 | 24.0 | 371 | 15907 | 8.86 | <0.01 |
| Left caudate, anterior | 15.8 | -20.0 | 10.0 | 94 | 4030 | 5.29 | <0.01 |
| Right putamen, insula | -36.8 | 11.5 | -0.5 | 125 | 5359 | 3.16 | <0.01 |
| $D2BP_{fally}$ x BMI *(b)* | | | | | | | |
| Right caudate | -12.2 | -9.5 | 3.0 | 36 | 1544 | 1.8 | <0.05 |
| Right thalamus | -1.8 | 8.0 | 6.5 | 28 | 1201 | -3.4 | >0.10 |
| Left caudate | 12.2 | -6.0 | 6.5 | 27 | 1158 | 3.7 | >0.10 |
| $D2BP_{raclo}$ x BMI *(c)* | | | | | | | |
| Left putamen | 29.8 | 8.0 | 3.0 | 172 | 7375 | -4.10 | <0.01 |
| Right putamen | -29.8 | 8.0 | -0.5 | 162 | 6946 | -4.16 | <0.01 |
| Voxelwise difference in $D2BP_{raclo}$ vs BMI and $D2BP_{fally}$ vs BMI correlation coefficients *(d)* | | | | | | | |
| Left caudate/putamen | 8.8 | 1.0 | 6.5 | 58 | 2487 | -0.63 | <0.01 |
| Right globus pallidus | -15.8 | 8.0 | 3.0 | 29 | 1243 | -0.64 | >0.10 |
| Right caudate/putamen | -12.2 | -6.0 | 3.0 | 26 | 1115 | -0.46 | >0.10 |

*a.* 1 sample t-test, n=54, $k_e$=20, $p_{uncorrected}$ = 0.01
*b.* 1 sample t-test, n=57, $k_e$=20, $p_{uncorrected}$ = 0.1
*c.* 1 sample t-test, n=56, $k_e$=20, $p_{uncorrected}$ = 0.1
*d.* Paired samples t-test, n=54, $k_e$=20, $p_{uncorrected}$ = 0.01
*e.* For (d), value reflects peak difference between correlation coefficients

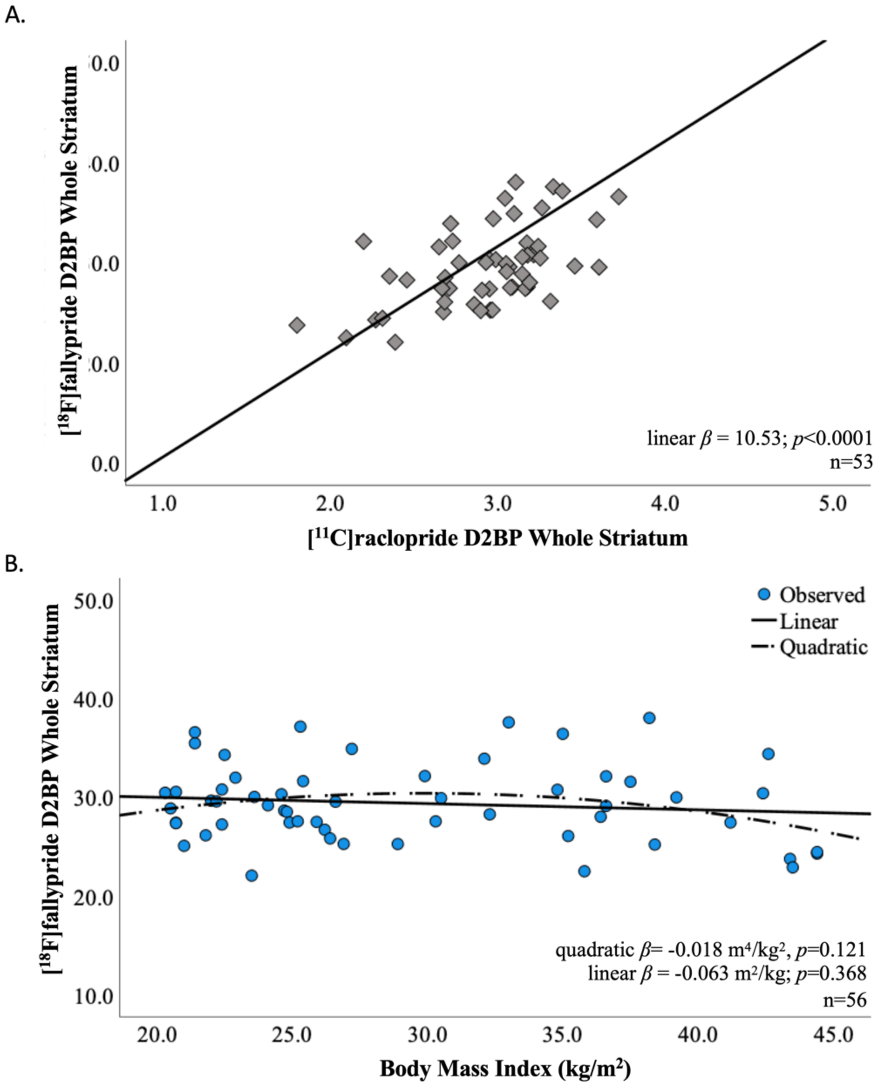


**Supplementary Figure 4. Re-analysis of primary aims exclusive of one outlier [$^{18}$F] fallypride datapoint.** Primary reported conclusions are robust to inclusion or exclusion of outlier. (A) Relationship between D2BP measurements using [$^{11}$C]raclopride and [$^{18}$F]fallypride excluding one outlier datapoint (n=53) (B) and relationship between D2BP measured by [$^{18}$F]fallypride and BMI (n=56).

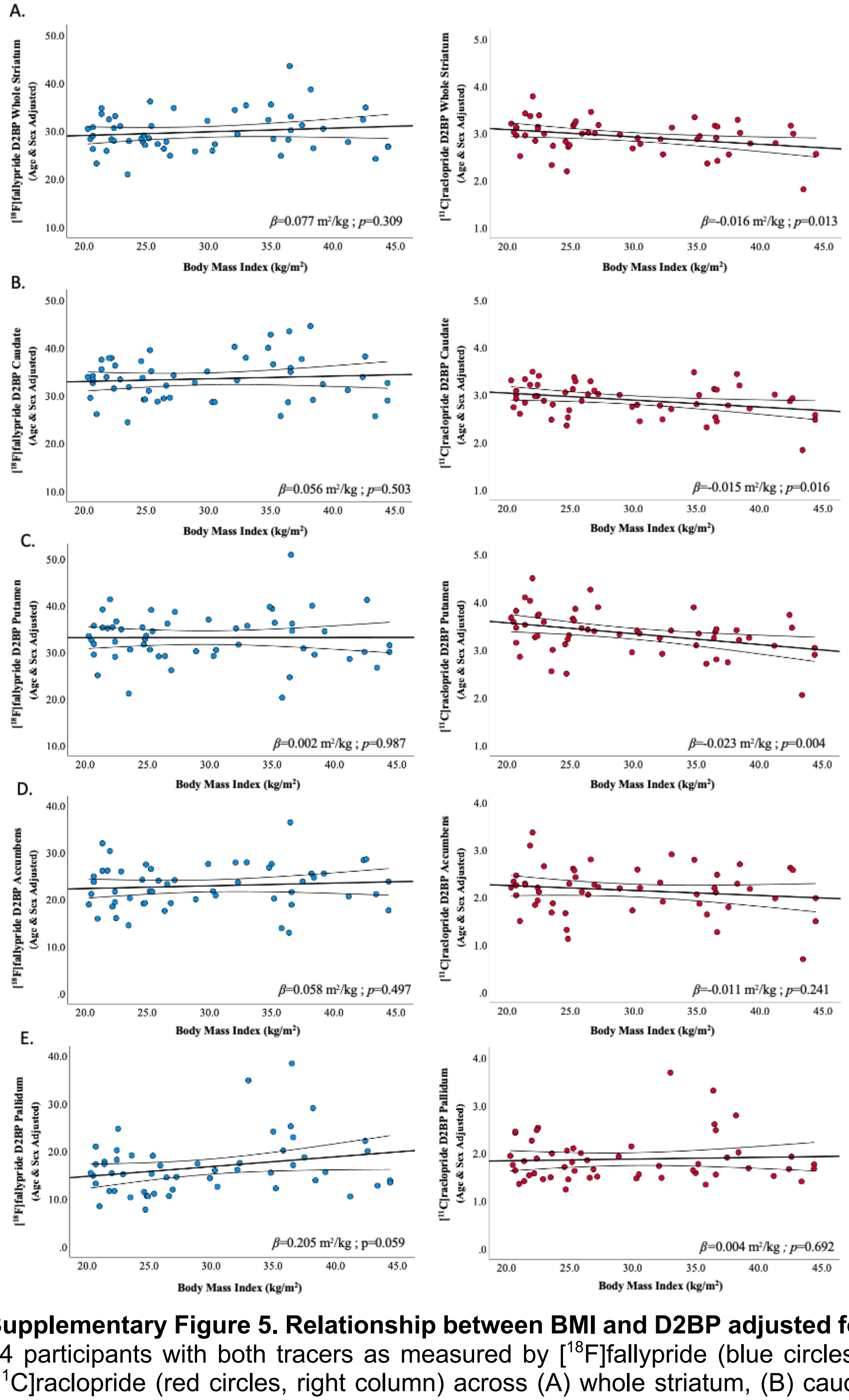


**Supplementary Figure 5. Relationship between BMI and D2BP adjusted for age and sex** for 54 participants with both tracers as measured by [18F]fallypride (blue circles, left column) and [11C]raclopride (red circles, right column) across (A) whole striatum, (B) caudate, (C) putamen, (D) accumbens, and (E) pallidum. Regression line and 95%CI indicated. Uncorrected for multiple comparisons.

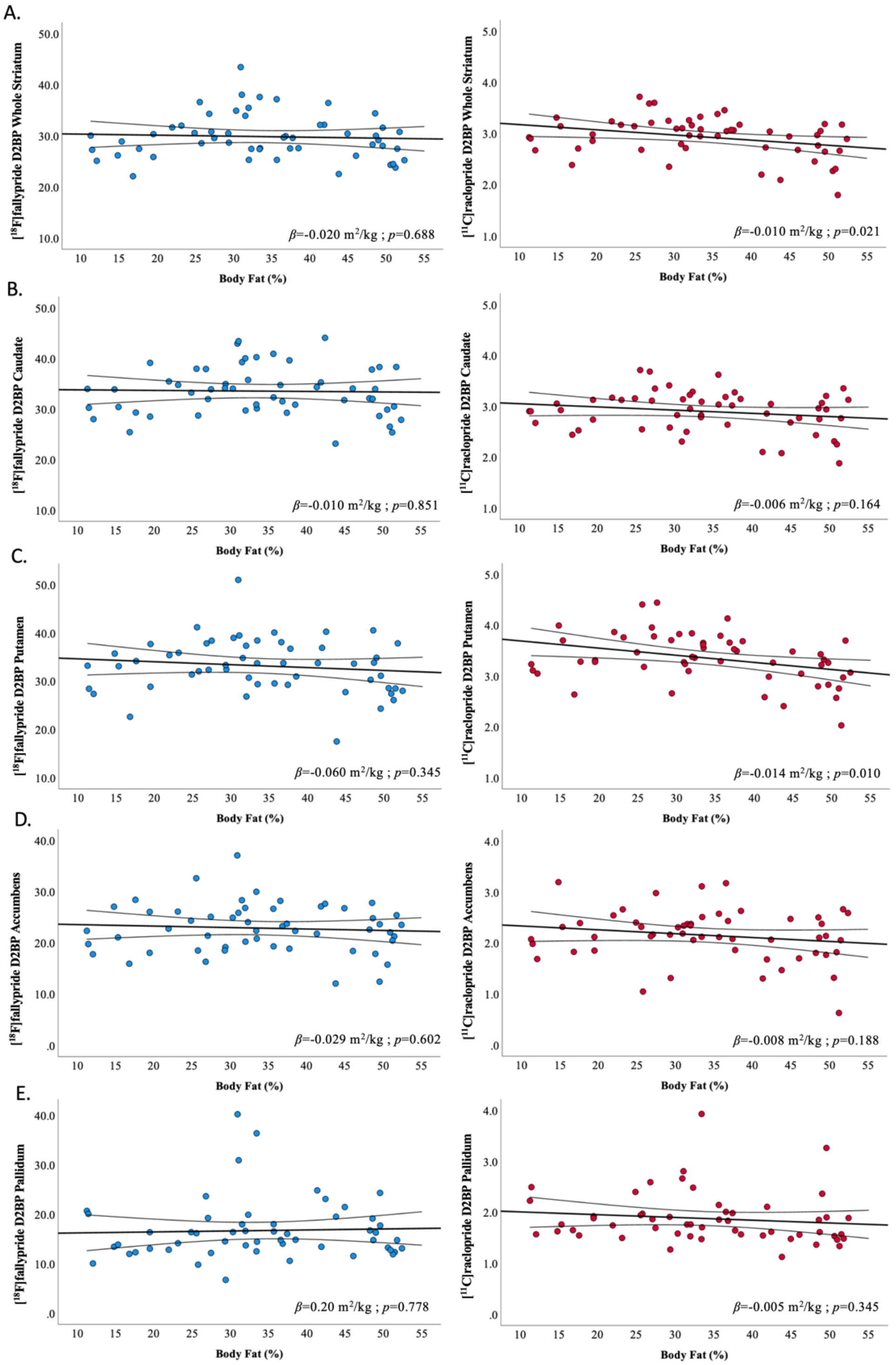


**Supplementary Figure 6. Relationship between percent body fat and D2BP** for 54 participants as measured by [$^{18}$F]fallypride (blue circles, left column) and [$^{11}$C]raclopride (red circles, right column) across (A) whole striatum, (B) caudate, (C) putamen, (D) accumbens, and (E) pallidum. Regression line and 95%CI indicated. Uncorrected for multiple comparisons.

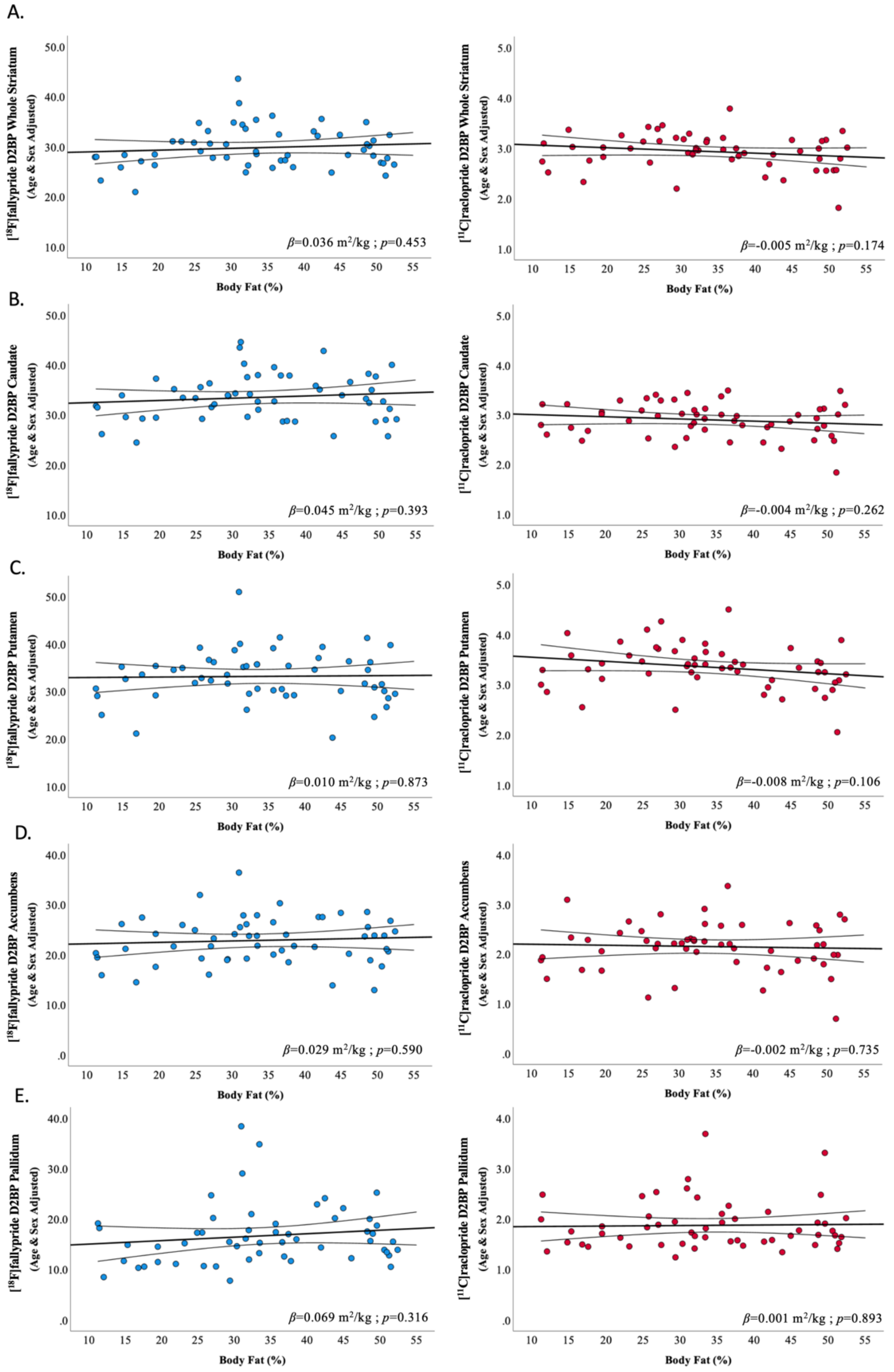


**Supplementary Figure 7. Relationship between percent body fat and D2BP adjusted for age and sex** for 54 participants as measured by [$^{18}$F]fallypride (blue circles, left column) and [$^{11}$C]raclopride (red circles, right column) across (A) whole striatum, (B) caudate, (C) putamen, (D) accumbens, and (E) pallidum. Regression line and 95%CI indicated. Uncorrected for multiple comparisons.

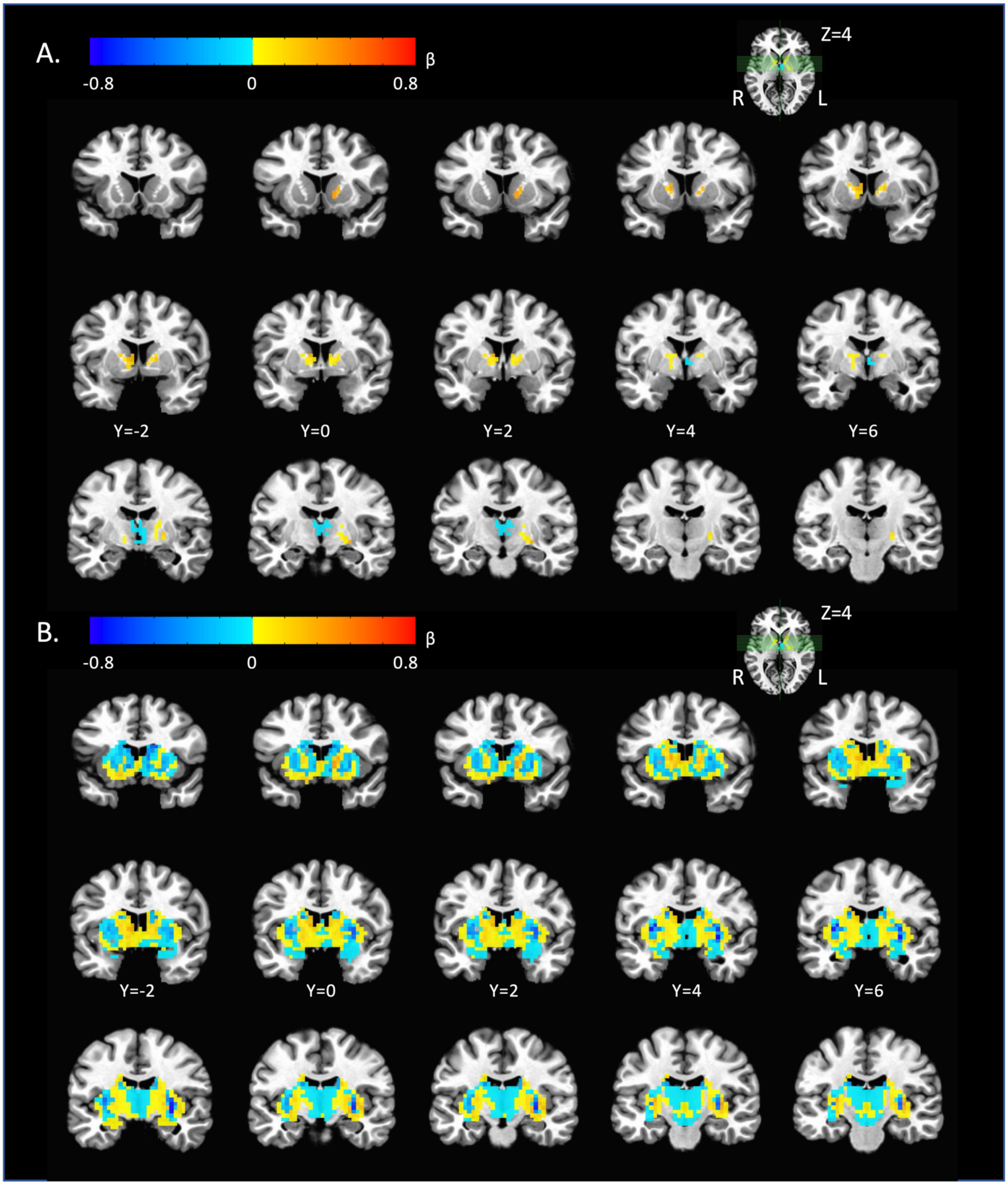


**Supplementary Figure 8. Voxelwise analysis of relationship between $D2BP_{fally}$ and BMI across n=57 participants.** (A) Thresholded (See Supplementary Table 1 for peak coordinates) (B) Unthresholded maps.

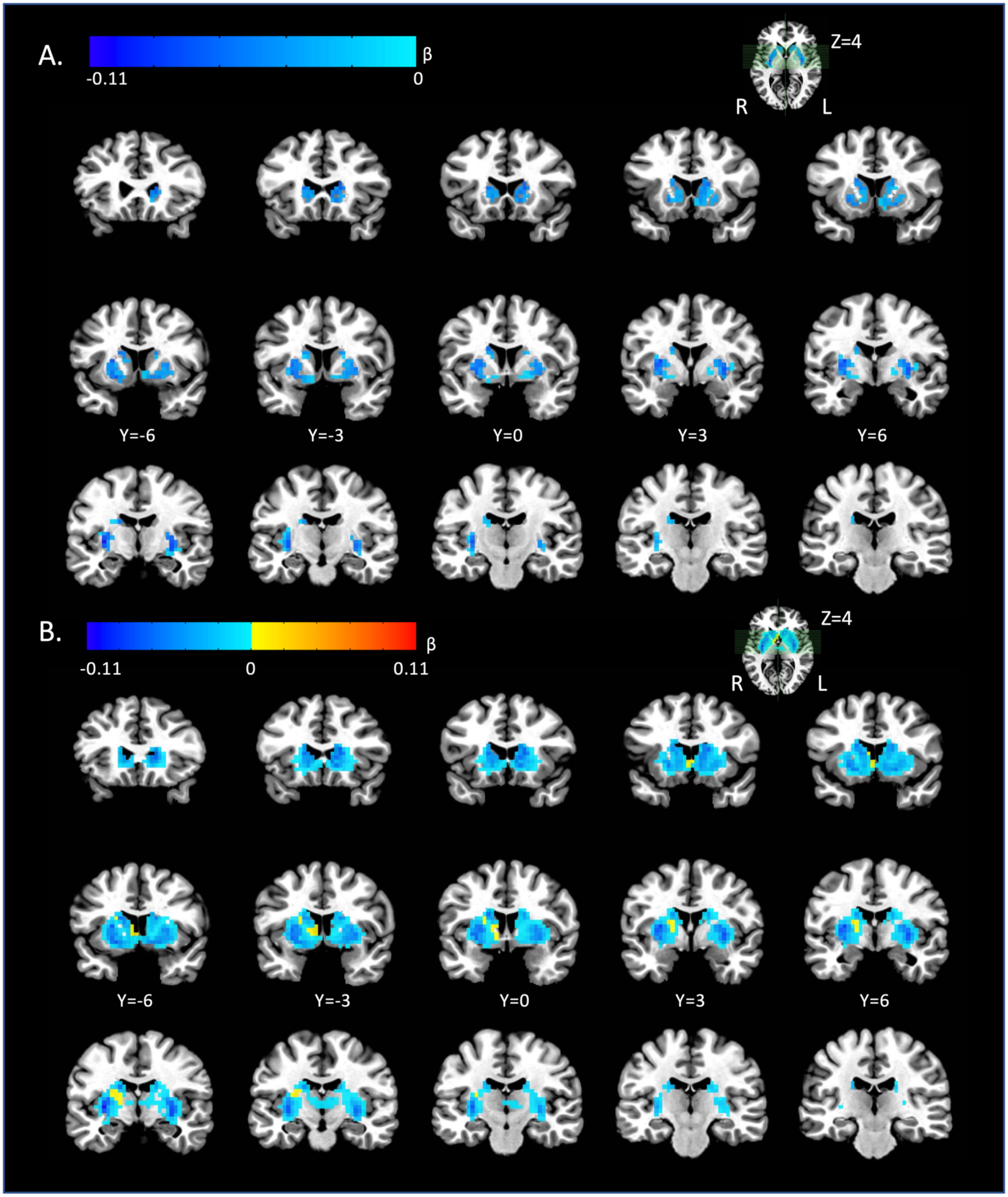


**Supplementary Figure 9. Voxelwise analysis of relationship between D2BP$_{raclo}$ and BMI** across n=56 participants. (A) Thresholded. (B) Unthresholded maps.

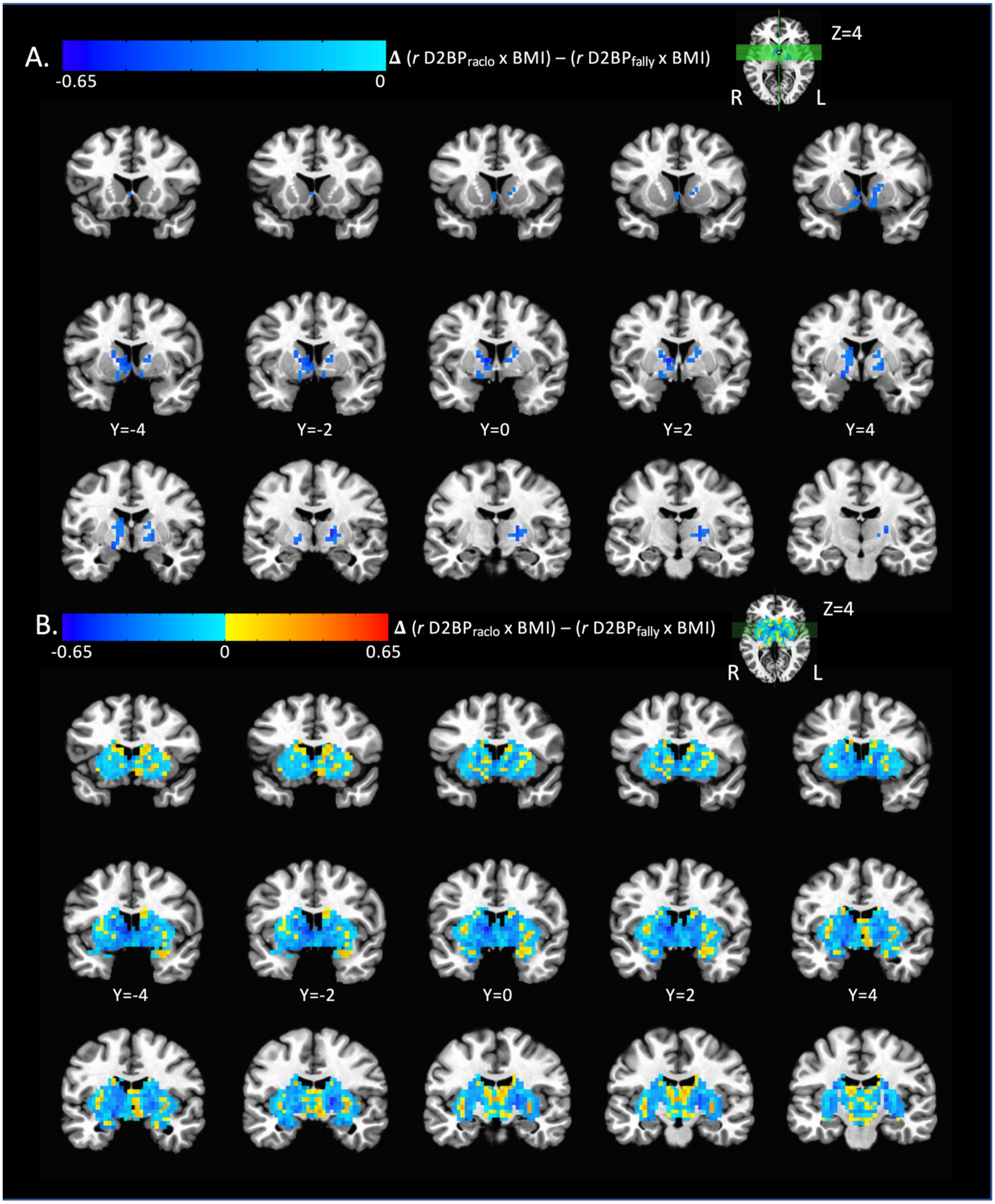


**Supplementary Figure 10. Voxelwise analysis of difference between normalized correlation coefficient for $D2BP_{raclo}$ x BMI and normalized correlation coefficient for $D2BP_{fally}$ x BMI** across n=54 participants. (A) Thresholded. (B) Unthresholded maps.
